# Psychosocial and Behavioral Responses and SARS-CoV-2 Transmission Prevention Behaviors while Working during the COVID-19 Pandemic

**DOI:** 10.1101/2021.05.11.21256774

**Authors:** Araliya M. Senerat, Zachary Pope, Sarah Rydell, Aidan Mullan, Veronique Roger, Mark A. Pereira

## Abstract

**Background:** The impact of coronavirus disease-2019 (COVID-19) on psychosocial and behavioral responses of the non-healthcare workforce is unknown. This study investigated these outcomes in this population during the pandemic while also evaluating transmission prevention behavior implementation at the workplace.

**Methods:** We deployed the baseline questionnaire of a prospective online survey from November 2020-February 2021 to U.S.-based employees. The survey included questions on psychosocial and behavioral responses in addition to transmission prevention behaviors (e.g., mask wear). Select questions asked employees to report perceptions and behaviors ‘before’ and ‘during’ the COVID-19 pandemic. Data were analyzed descriptively and stratified by work from home (WFH) percentage.

**Results:** In total, 3,607 employees completed the survey from eight companies. Most participants (70.0%) averaged ≥90% of their time WFH during the pandemic. Employees reported increases in stress (54.0%), anxiety (57.4%), fatigue (51.6%), and feeling unsafe (50.4%) from before to during the pandemic, while feeling a lack of companionship (60.5%) and isolation from others (69.3%). Productivity was perceived to decrease, and non-work screen time and alcohol consumption to increase, for 43.0%, 50.7%, and 25.1% of employees, respectively, from before to during the pandemic. Adverse changes were worse among those with lower WFH percentages. Most employees reported wearing a mask (98.2%), washing hands regularly (95.7%), and physically distancing (93.6%) when at workplace.

**Conclusion:** Results suggested worsened psychosocial and behavioral outcomes from before to during the COVID-19 pandemic and higher transmission prevention behavior implementation among non-healthcare employees. Observations provide novel insight into how the COVID-19 pandemic has impacted non-healthcare employees.

## Introduction

Coronavirus disease-19 (COVID-19), caused by the severe acute respiratory syndrome coronavirus-2 (SARS-CoV-2) virus, has resulted in high morbidity and mortality since being declared a global pandemic in March 2020.^1-3^ As most SARS-CoV-2 infections are transmitted by asymptomatic individuals,^4^ infection control measures have been and continue to be vital even as effective vaccines are administered. Across many workforce sectors, companies have implemented mitigation strategies, including mandates that employees work from home if possible, while also requiring engagement in SARS-CoV-2 transmission prevention behaviors (e.g., physical distancing, mask wear, hand/surface disinfection).^5-7^ While these mitigation strategies may help reduce COVID-19 infection rates, unintended negative consequences on employees may arise (e.g., feelings of isolation, stress/anxiety, adverse health behavior changes).

The COVID-19 pandemic has led to increased prevalence^8-14^ of higher self-reported stress, anxiety, depression, episodes of acute panic, obsessive behaviors, and post-traumatic stress syndrome (PTSD), among others.^10^ Widespread quarantine and fear of self/loved ones getting COVID-19 are among the largest contributors to observed poorer psychosocial health.^10,11^ Mental health problems are also higher among those diagnosed with COVID-19 compared to those not directly affected,^15^ as well as among those who quarantine to prevent the spread of COVID-19 and other diseases.^16^ Within the workplace, past literature noted that being a healthcare worker or caregiver of persons with SARS-CoV in 2003 was associated with higher likelihood of negative psychosocial outcomes, exacerbating the impact of the COVID-19 pandemic on this employee cohort.^10^ How past infectious diseases and the COVID-19 pandemic have impacted the psychosocial health of non-healthcare employees has scantly been studied.

The SARS-CoV-2 virus spreads primarily via small airborne infectious particles (i.e., aerosols) that infected individuals may generate when breathing, coughing, sneezing, or talking.^17-27^ A less common transmission route is via larger respiratory droplets that deposit onto nearby surfaces.^25,27-52^ A recent meta-analysis^53^ and several standalone investigations^54-59^ have shown the ability of mask wearing, physical distancing, and hand/surface disinfection to mitigate transmission of various coronaviruses. Thus, SARS-CoV-2 prevention behaviors in the workplace are critical to reduce COVID-19 infection rates, with engagement in these behaviors also associated with lower levels of stress, anxiety, and depression.^11^

Governmental recommendations are in place for SARS-CoV-2 transmission prevention at the workplace.^60^ Yet, few data exist across non-healthcare employee workforce sectors regarding employee: (1) psychosocial outcomes (e.g., stress, anxiety, safety) and perceptions as it relates to working during the COVID-19 pandemic; (2) behavioral outcomes (e.g., smoking, alcohol consumption, physical activity); and (3) implementation of SARS-CoV-2 transmission prevention behaviors at the workplace. Survey collection of such data would inform workplace strategies to reduce negative psychosocial outcomes among employees as more individuals return regularly to the workplace, while also offering insight on workplace mitigation of COVID-19 and other respiratory illnesses.

The purpose of our study was to examine COVID-19-related psychosocial outcomes, prevention practices, and health behaviors among employees across numerous workforce sectors.

## Methods

This study was approved by the University of Minnesota (IRB #: STUDY00010426) and Mayo Clinic (IRB #: 20-007642) Institutional Review Boards. All study participants gave informed consent.

### Study Design and Survey Development

We designed a prospective online survey entitled “*<u>C</u>haracterizing <u>A</u>wareness of SARS-CoV-2 <u>P</u>reven<u>T</u>ion and <u>U</u>nderstanding <u>R</u>esponses and <u>E</u>xperiences (CAPTURE) Survey*”, with this survey deployed at baseline, three months, six months, and one year. As the follow-up surveys are still ongoing, only the baseline results are presented here. The CAPTURE Survey consisted of 48 questions across eight sections. These sections included consent/screening, demographics, job description, and general health-related questions as well as sections regarding: (1) the socioeconomic impact of COVID-19; (2) personal SARS-CoV-2 prevention behaviors at the workplace; (3) perceptions of the importance/efficacy of SARS-CoV-2 prevention behaviors; (4) workplace COVID-19 culture and practices; (5) psychosocial experiences and perceptions before and during the COVID-19 pandemic; and (6) health behaviors before and during the COVID-19 pandemic. See ‘Measures’ and Appendix A for description of survey components/questions. We piloted the CAPTURE Survey twice with experts at the Well Living Lab, Mayo Clinic, and the University of Minnesota over 1.5 months before deployment. The CAPTURE Survey completion time was ∼12 minutes.

The CAPTURE Survey was deployed using the Mayo Clinic’s Qualtrics Platform. Survey responses were deidentified and given a unique study ID that consisted of a series of numbers that had no meaning to employees’ personal identifying characteristics. We used this ID to track each individual employee in the research database.

### Inclusion and Exclusion Criteria

Company-level inclusion criteria were: (1) any public or private company; and (2) in-business at time of baseline survey distribution. We assessed these company-level criteria before contacting each company regarding their employees’ potential participation in the CAPTURE Survey. Employee-level inclusion criteria were: (1) ≥18 years old; (2) English speaking; (3) currently employed by the company; and (4) working ≥50% of their workweek indoors given that SARS-CoV-2 is most likely to spread indoors. Employees were excluded if they were: (1) <18 years old; (2) unemployed; (3) working primarily outdoors; and (4) did not work within the U.S. We excluded employees working outside the U.S. given the differing cultures, practices, and severity of the COVID-19 pandemic between the U.S. and other countries.

### Measures

#### Consent/Screening, Demographics, Job Descriptions, and General Health

We gathered each employee’s age, employment status, employer (i.e., company), and U.S. location. Average percentage of workweek time working indoors was assessed on a sliding 0-100% scale in 1%-increments. Demographic information, job descriptions, and general health-related information were queried using questions adapted from the *Stand and Move at Work* (SMW) group randomized trial and the Coronary Artery Risk Development in Young Adults (CARDIA) study.^61,62^

#### Socioeconomic Impact of COVID-19

We developed questions assessing the average number of hours/week that the employee worked in total and face-to-face at the workplace. We assessed average percentage of workweek hours spent working from home (WFH) on a sliding 0-100% scale in 1%-increments, with this question asked in parallel to acquire ‘before’ and ‘during’ COVID-19 WFH percentages. We also adapted questions from the SMW trial and the Environmental Influences on Child Health Outcomes (ECHO) study to examine health insurance coverage and doctors visits differed before and during COVID-19.^62,63^

#### Psychosocial Outcomes and Perceptions

We assessed employee’s frequency of feeling stressed, anxious, fatigued, and unsafe before and during COVID-19 on a 5-point Likert-type scale (1: *Never*; 5: *All the time*), while also using a 5-point Likert-type scale (1: *Low*; 5: *High*) to assess employees’ perceived productivity before and during COVID-19. These questions were adapted from the SMW trial.^62^ We developed questions to evaluate whether employees felt threatened or afraid of COVID-19, as well as their fear of catching the disease when around other people, assessed on a 5-point Likert-type scale (1: *Strongly Disagree*; 5: *Strongly Agree*). We adapted questions from the SMW trial to assess employee engagement in health behaviors before and during COVID-19. These health behaviors included physical activity, non-work-related screen time, sleep, and alcohol consumption.

#### SARS-CoV-2 Prevention Behaviors at the Workplace

We included questions that asked about frequency of: (1) SARS-CoV-2 prevention behaviors (mask wear, glove wear, handwashing, physical distancing, surface disinfection) that an employee and their coworkers have taken when at the workplace; and (2) company provision of SARS-CoV-2 prevention supplies. These questions were asked on a 5-point Likert-type scale (1: *Never*; 5: *Always*). We also assessed what types of SARS-CoV-2 prevention behavior training had been provided by employees’ companies and whether the employee perceived SARS-CoV-2 prevention behaviors to be effective at preventing the spread of COVID-19, the latter on a 4-point Likert-type scale (1: *Not Important*; 4: *Very Important*). Importantly, these questions had “not applicable” and/or “prefer not to answer” options given the sensitivity of these questions and the larger social discussion around COVID-19 in the U.S.

#### COVID-19 Symptomology and Diagnosis

We inquired whether the employee had experienced any COVID-19 symptoms and/or been diagnosed with COVID-19 using questions adapted from ECHO, Epidemic-Pandemic Impacts Inventory, CARDIA, Three-Item Loneliness Scale, and Coronavirus Questionnaire.^61-66^

### Recruitment & Survey Deployment

We contacted a convenience sample of 234 U.S. companies regarding CAPTURE Survey participation through word of mouth, emails, LinkedIn messages, and website form submissions. Figure 1 reviews the number of companies contacted and interested as well as the number of participating companies. We spoke with human resource personnel, managers, and/or supervisors (hereafter, key contacts) within companies returning correspondence. A short, informational presentation was provided to key contacts of interested companies that outlined the CAPTURE Survey’s aims, company and employee participation requirements, and the potential benefits of the company’s participation. Companies were given two CAPTURE Survey deployment choices if they agreed to have employees participate: (1) company sent out a company-wide email with CAPTURE Survey link and informational materials that invited employee participation; or (2) company collected email addresses of interested employees and shared those email addresses with us after which we deployed the CAPTURE Survey link to those email addresses only. Four companies chose to generate their own company-wide deployment email, and we sent out company-provided emails to four companies. While employees from participating companies were strongly encouraged to participate, they were told participation was completely voluntary and had no effect on their company employment or relationship with us. The CAPTURE Survey was sent to employees of participating companies starting November 20, 2020, and the baseline survey closed February 8, 2021. A reminder email was sent approximately 5-7 business days after initial deployment with each company, with the survey open to employees for up to three weeks after the initial deployment date.

**Figure 1.**
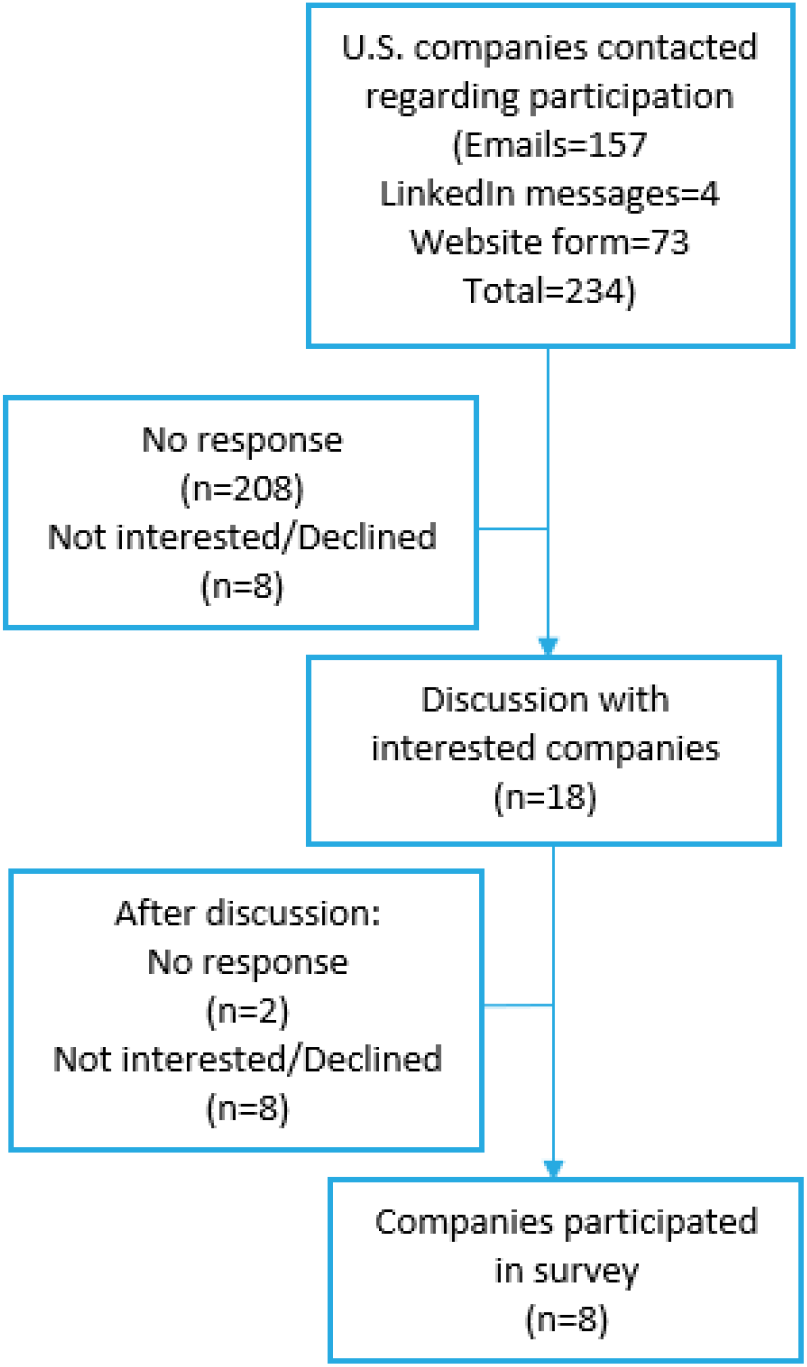
Flowchart of the companies who expressed interest and participation status.

### Statistical Analyses

CAPTURE Survey responses were downloaded from Mayo Clinic’s Qualtrics Platform. Data were cleaned using Microsoft Excel and uploaded to Python 3.9 for further cleaning and analyses using Jupyter Notebook for Windows 1.3.1093. Duplicates were examined by the date of first survey completion and CAPTURE Survey progress completion for each duplicate entry. Of the duplicates, we kept the CAPTURE Survey entry with the highest progress completion as long as survey completion fell within a 3-week time period allocated for employees of a given company to complete the survey.

In addition to determining prevalence and mean responses for the entire cohort, we also stratified by WFH categories: ≤25%, 26-50%; 51-75%; >75%. Further, we compared employee responses to questions asking about psychosocial experiences, perceptions, and health behaviors ‘before’ and ‘during’ the COVID-19 pandemic. Likert responses were coded numerically and the mean change between responses ‘before’ and ‘during’ the pandemic was calculated for the whole sample as well as by WFH categories with 95% confidence intervals.

## Results

A total of 3,607 employees completed the CAPTURE survey from eight separate companies across the U.S. Seven companies were classified as industry, and one company was an academic institution. The overall mean response rate from employees within the eight companies was 14.9%, with intra-company response rates ranging from 6.8% to 63.8%. When data were stratified by company type, results were not materially different.

### Demographics

Demographic results are observed in Table 1. Most employees were from the Midwest (83.5%). Mean age was 44.7 ±12.1 years, with most employees classified as ‘Professionals’ (49.5%), followed by ‘Executive, Administrator, or Senior Manager’ (13.5%), and ‘Clerical and Administrative Support’ (11.8%). Perceived health was in ‘Good’, ‘Very Good’, or ‘Excellent’ condition for most (91.7%). The percentage of employees reporting that their doctor had told them they had a medical condition ranged from 2.9% for heart disease and stroke (combined) to 24.0% for mood disorder. Most employees (62.1%) reported reduced in-person contact with family who live outside the home, friends, colleagues at work, and events in the community because of COVID-19.

**Table 1.** Demographic results of the Baseline CAPTURE Survey.^a^

| **Gender** | **Count** | **%** |
| --- | --- | --- |
| Female/Women | 2292 | 67.4% |
| Male/Men | 1063 | 31.3% |
| Other/Nonbinary | 33 | 1.0% |
| **Age** |  |  |
| <35 | 896 | 26.9% |
| 35-65 | 2309 | 71.8% |
| >65 | 125 | 3.8% |
| **Hispanic or Latino/Latina/Latinx** |  |  |
| Yes | 124 | 3.5% |
| **Which of the following best describes you?** |  |  |
| Asian, Black or African-American | 217 | 6.4% |
| Other | 216 | 6.4% |
| White | 2924 | 86.0% |
| **Current marital status** |  |  |
| Married or partnered | 2469 | 72.6% |
| **Highest level of schooling completed** |  |  |
| Less than Bachelor's degree | 364 | 10.7% |
| Bachelor's degree | 1260 | 37.1% |
| Master's, Professional, or Doctoral degree | 1762 | 51.8% |

<sup>a</sup> Prefer not to answer were not included in the above table and were less than 2% of the responses.

### Socioeconomic Impact of COVID-19

Most (85.7%) employees stated they spent >90% of their time indoors when working, 90.5% stated they spent an average of 33-60 hours per week working, and 58.8% stated they currently had no face-to-face, in-person interactions with either coworkers or the public while completing their job-related duties. Few employees (0.2%) reported being furloughed or furloughed previously (or temporarily laid off). Approximately 68% believed COVID-19 had a negative impact on their work. Few (0.7%) lost health insurance or other coverage for medical care. As shown in Figure 2, with WFH percentage grouped into the four 25% categories, the distribution shifts dramatically from ‘before’ to ‘during’ the COVID-19 pandemic.

**Figure 2.**
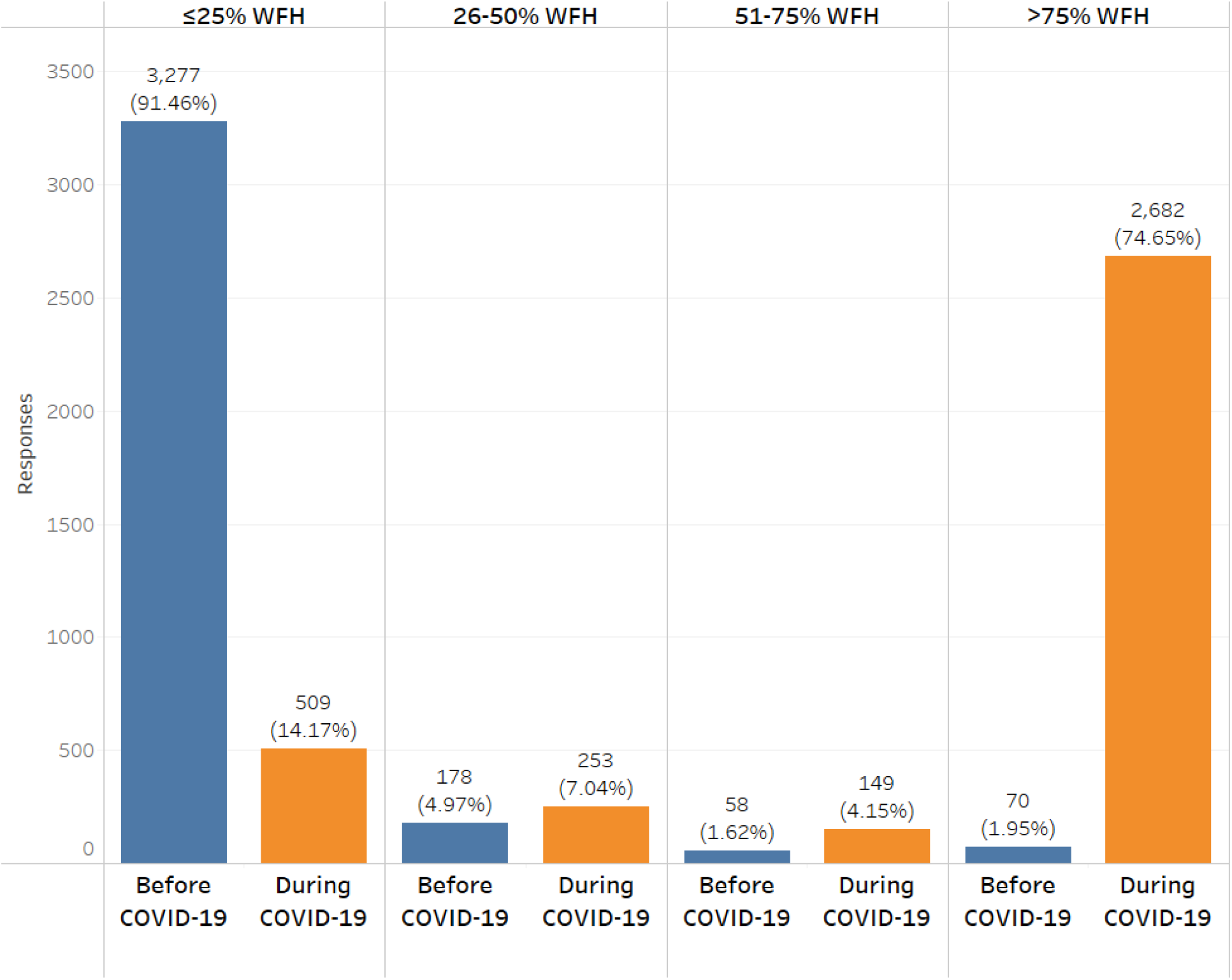
WFH Categories Before & During the COVID-19 Pandemic

### Psychosocial Outcomes and Perceptions

During the pandemic, many employees reported feeling stressed (53.1%), anxious (43.8%), and fatigued (41.1%) ‘Quite a bit’ or ‘All the time’. The prevalence of employees agreeing they felt threatened about COVID-19 or afraid of COVID-19 were 39.4% and 60.0%, respectively, while 60.3% reported being stressed around other people because they worried they would catch COVID-19. In total, 32.4% reported being ‘Moderately’ or ‘Very Worried’ about contracting COVID-19 while at work, and 34.1% reported being ‘Moderately’ or ‘Very Worried’ about being an asymptomatic carrier.

Table 2 shows the changes in psychosocial and behavioral responses from before to during the COVID-19 pandemic, with mean change and 95% confidence intervals for each parameter. More than half of employees reported an increase in stress, anxiety, fatigue, feeling unsafe, a lack of companionship, and a feeling of being isolated from others. For the behavioral outcomes, a high percentage of employees reported a decrease in productivity and physical activity, while increases were observed for non-work screen time, sleep, and alcohol consumption.

**Table 2.**
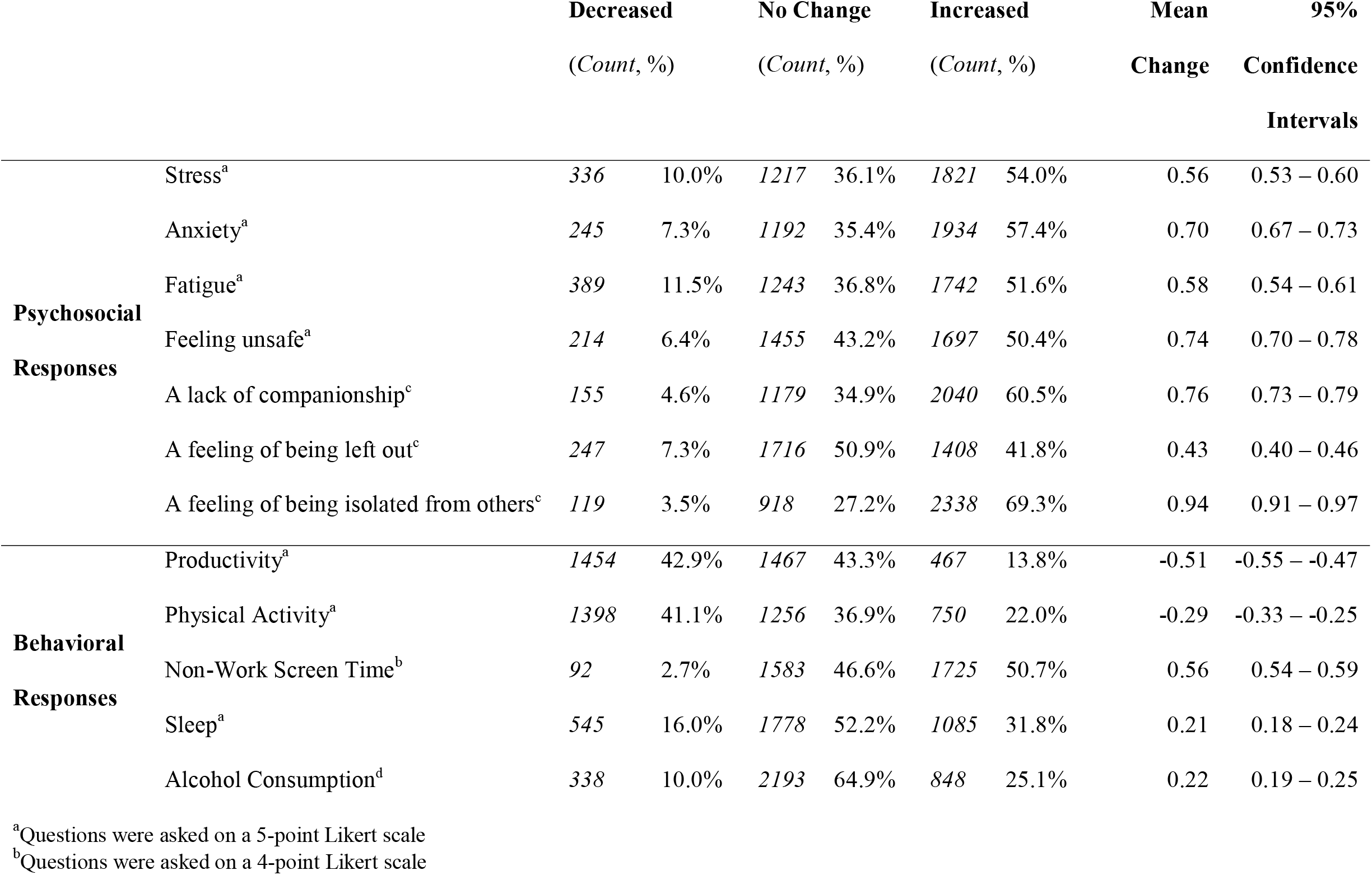
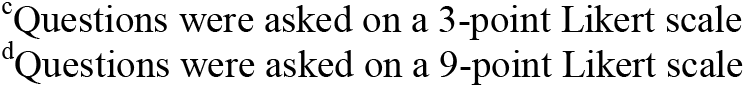
Psychosocial & Behavioral Changes from Before to During the COVID-19 Pandemic

|  |  | **Decreased** |  | **No Change** |  | **Increased** |  | **Mean** | **95%** |
| --- | --- | --- | --- | --- | --- | --- | --- | --- | --- |
|  |  | *(Count, %)* |  | *(Count, %)* |  | *(Count, %)* |  | **Change** | **Confidence** |
|  |  |  |  |  |  |  |  |  | **Intervals** |
| **Psychosocial Responses** | Stress<sup>a</sup> | 336 | 10.0% | 1217 | 36.1% | 1821 | 54.0% | 0.56 | 0.53 – 0.60 |
|  | Anxiety<sup>a</sup> | 245 | 7.3% | 1192 | 35.4% | 1934 | 57.4% | 0.70 | 0.67 – 0.73 |
|  | Fatigue<sup>a</sup> | 389 | 11.5% | 1243 | 36.8% | 1742 | 51.6% | 0.58 | 0.54 – 0.61 |
|  | Feeling unsafe<sup>a</sup> | 214 | 6.4% | 1455 | 43.2% | 1697 | 50.4% | 0.74 | 0.70 – 0.78 |
|  | A lack of companionship<sup>c</sup> | 155 | 4.6% | 1179 | 34.9% | 2040 | 60.5% | 0.76 | 0.73 – 0.79 |
|  | A feeling of being left out<sup>c</sup> | 247 | 7.3% | 1716 | 50.9% | 1408 | 41.8% | 0.43 | 0.40 – 0.46 |
|  | A feeling of being isolated from others<sup>c</sup> | 119 | 3.5% | 918 | 27.2% | 2338 | 69.3% | 0.94 | 0.91 – 0.97 |
| **Behavioral Responses** | Productivity<sup>a</sup> | 1454 | 42.9% | 1467 | 43.3% | 467 | 13.8% | -0.51 | -0.55 – -0.47 |
|  | Physical Activity<sup>a</sup> | 1398 | 41.1% | 1256 | 36.9% | 750 | 22.0% | -0.29 | -0.33 – -0.25 |
|  | Non-Work Screen Time<sup>b</sup> | 92 | 2.7% | 1583 | 46.6% | 1725 | 50.7% | 0.56 | 0.54 – 0.59 |
|  | Sleep<sup>a</sup> | 545 | 16.0% | 1778 | 52.2% | 1085 | 31.8% | 0.21 | 0.18 – 0.24 |
|  | Alcohol Consumption<sup>d</sup> | 338 | 10.0% | 2193 | 64.9% | 848 | 25.1% | 0.22 | 0.19 – 0.25 |

<sup>a</sup>Questions were asked on a 5-point Likert scale
<sup>b</sup>Questions were asked on a 4-point Likert scale
<sup>c</sup>Questions were asked on a 3-point Likert scale
<sup>d</sup>Questions were asked on a 9-point Likert scale

Figure 3 shows the mean change and 95% confidence intervals for the psychosocial and behavioral responses stratified by WFH categories of >75% v. ≤25% of worktime. As shown, mean increases were higher in the ≤25% WFH group for stress, anxiety, fatigue, and feeling unsafe. Decreases in productivity were larger for WFH ≤25%, while an increase in sleep was only observed in WFH >75%.

**Figure 3.**
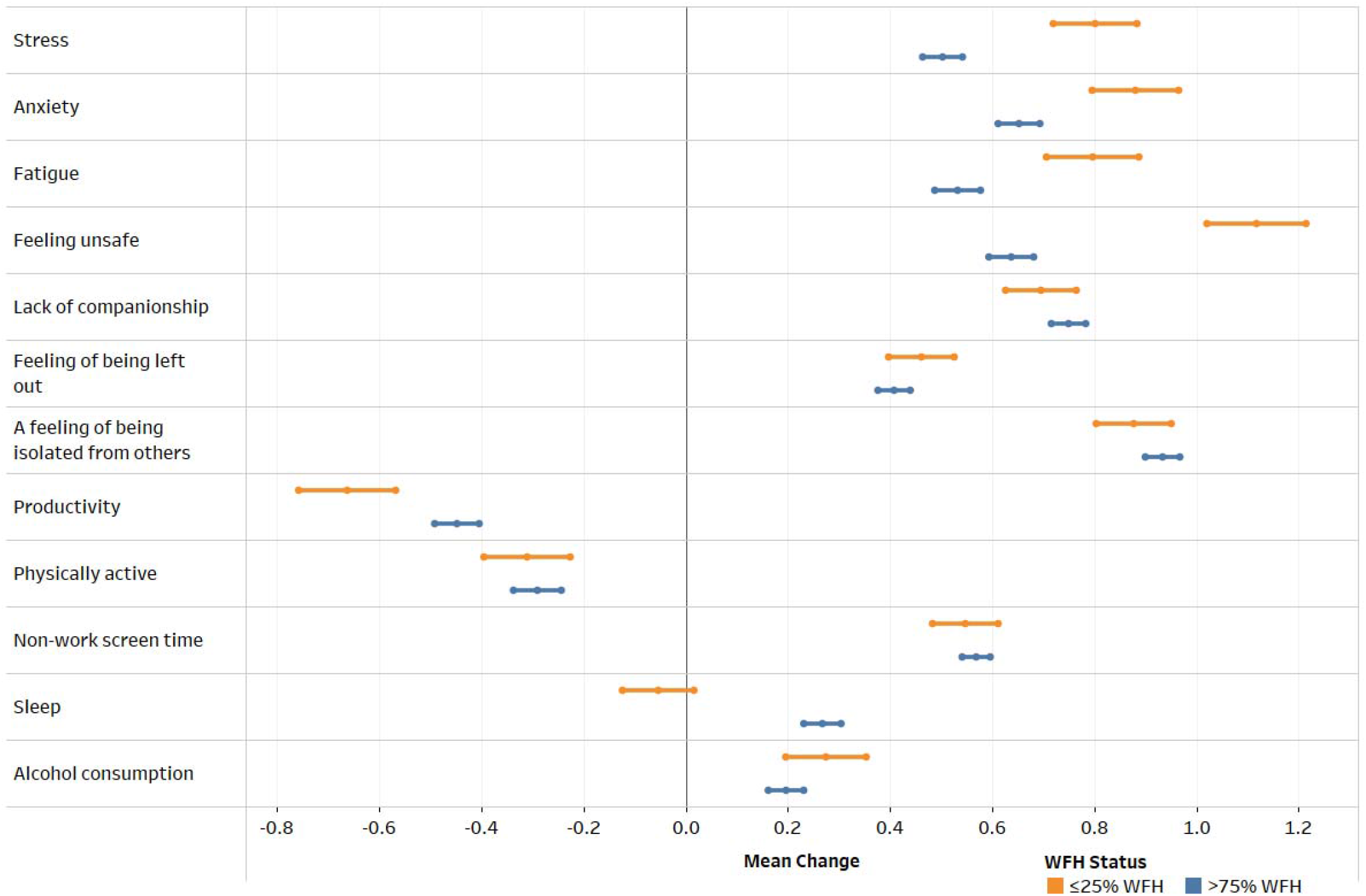
Psychosocial & Behavioral Responses (Mean, 95% Confidence Intervals) from Before to During the COVID-19 Pandemic Stratified by WFH Status (≤25% WFH vs >75% WFH)

### SARS-CoV-2 Prevention Behaviors at the Workplace

Employees reported ‘Often’ or ‘Always’ engaging in personal SARS-CoV-2 prevention behaviors at the workplace, as seen in Table 3. Glove wearing was an exception, with most reporting ‘Never’ or ‘Rarely’. Table 4 shows PPE, instructions, and practices provided and promoted by the employers. Most employees reported their employer provided web training (33.4%), reading materials (24.1%), or both (25.5%).

**Table 3.**
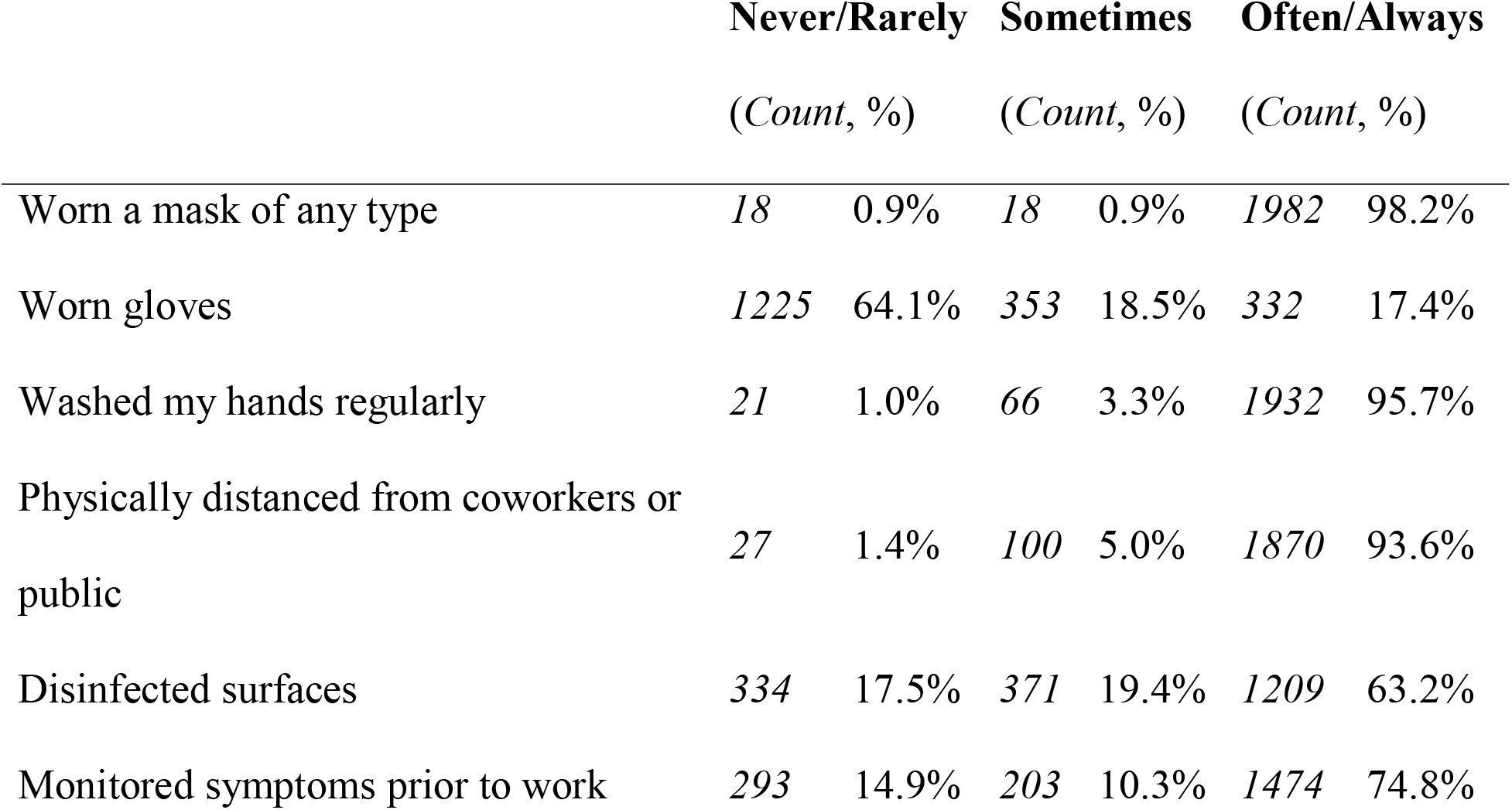
Personal COVID-19 Prevention Behaviors During COVID-19 Pandemic

|  | **Never/Rarely** |  | **Sometimes** |  | **Often/Always** |  |
| --- | --- | --- | --- | --- | --- | --- |
|  | *(Count, %)* |  | *(Count, %)* |  | *(Count, %)* |  |
| Worn a mask of any type | 18 | 0.9% | 18 | 0.9% | 1982 | 98.2% |
| Worn gloves | 1225 | 64.1% | 353 | 18.5% | 332 | 17.4% |
| Washed my hands regularly | 21 | 1.0% | 66 | 3.3% | 1932 | 95.7% |
| Physically distanced from coworkers or public | 27 | 1.4% | 100 | 5.0% | 1870 | 93.6% |
| Disinfected surfaces | 334 | 17.5% | 371 | 19.4% | 1209 | 63.2% |
| Monitored symptoms prior to work | 293 | 14.9% | 203 | 10.3% | 1474 | 74.8% |

**Table 4.**
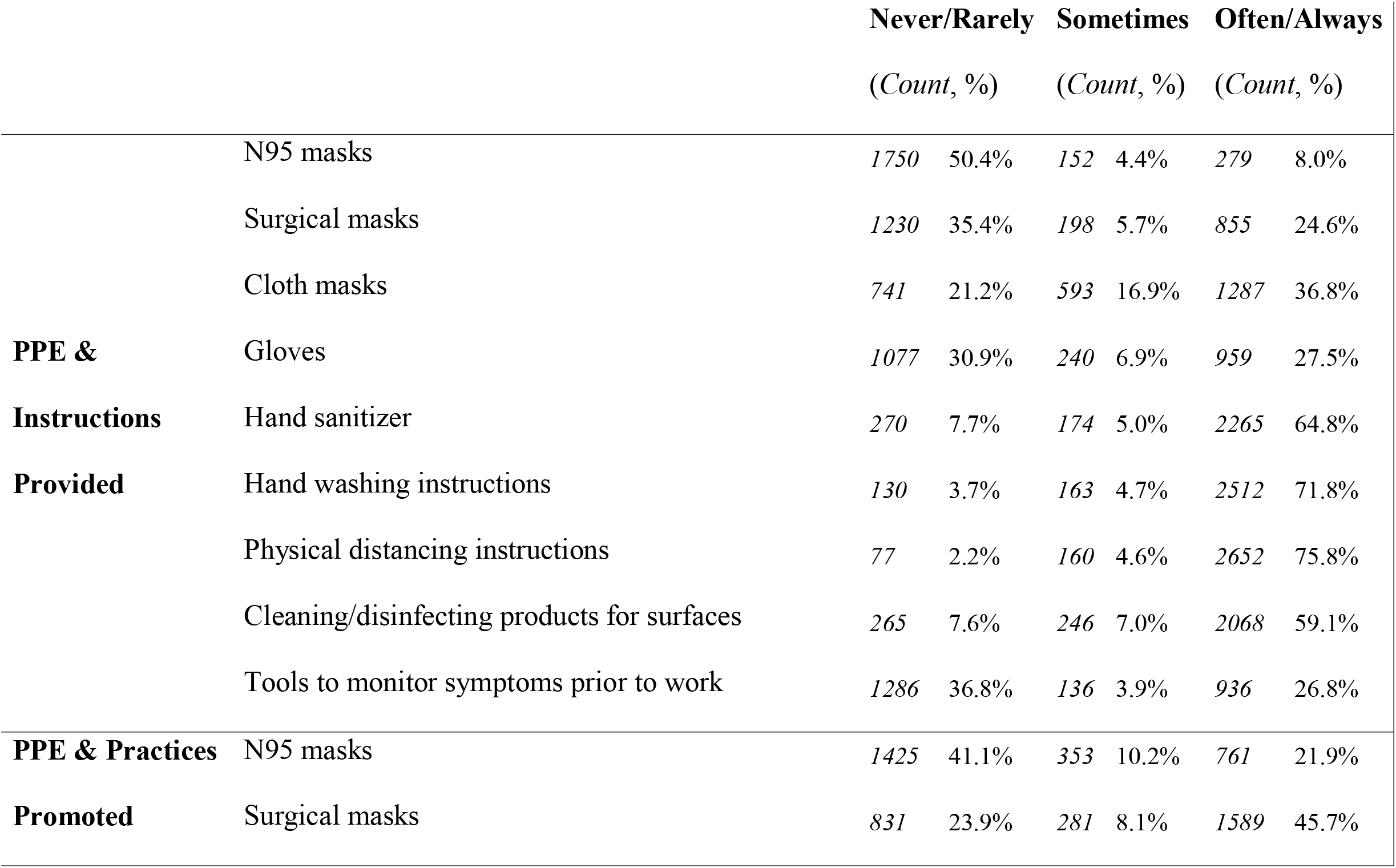
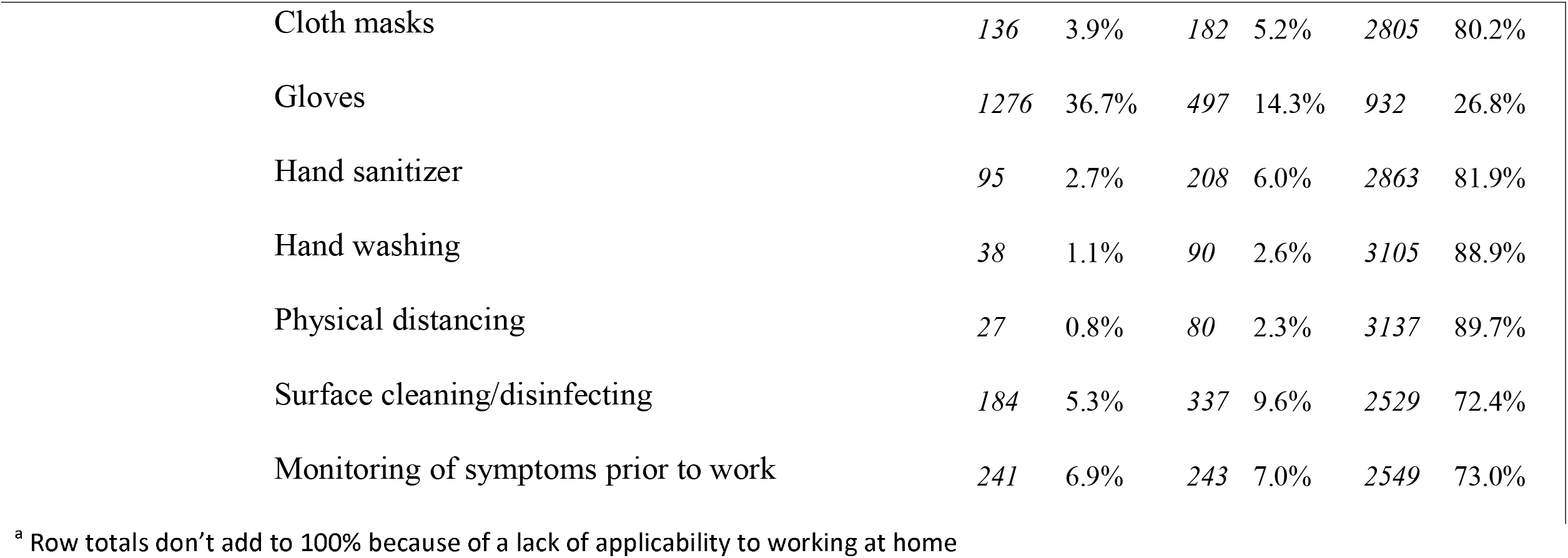
Workplace Culture, Practices, & PPE Provided & Promoted by Companies^a^

When asked about perceptions of the importance of specific prevention behaviors and practices, most employees reported wearing a mask, handwashing, and physical distancing to be ‘Very Important’ (93.3%, 89.1%, and 93.5%, respectively). Disinfecting surfaces was reported as ‘Very Important’ by some (45.5%), while most employees reported that wearing gloves was ‘Not Important’ (36.2%).

### COVID-19 Symptomology and Diagnosis

COVID-19 symptoms reported as lasting several hours more than usual were headaches (34.3%), followed by unusual fatigue (27.1%), malaise or general feeling of illness, discomfort or uneasiness (23.2%), and muscle aches (22.3%). In total, 13.1% of employees suspected they had a COVID-19 infection or had COVID-19 symptoms. Most reported having a negative nasal swab or spit test (52.9%) and a negative blood/antibody test (9.9%). Few had a positive nasal swab or spit test (4.0%) and positive blood/antibody test (1.0%).

## Discussion

We observed large increases in stress, anxiety, fatigue, and feeling unsafe among non-healthcare employees due to the COVID-19 pandemic. We also noted an increase in lack of companionship, feeling left out, or isolation when completing their work-related duties, with decreased productivity and physical activity reported. These changes may have been due to quarantining and the sudden shift to WFH. Most employees were afraid of COVID-19 and stressed about acquiring the disease in general. Interestingly, however, not as many employees were worried about contracting COVID-19 at work or being an asymptomatic carrier of COVID-19 while at work, regardless of WFH status. A high percentage of employees believed wearing a mask, handwashing, and physical distancing were important in preventing the spread of COVID-19, whereas wearing gloves and disinfecting surfaces were thought to be less important. While productivity and physical activity decreased for many, non-work screen time increased. Sleep also increased in a small percentage of employees and, in particular, those engaged in >75% WFH each week, which may be due to the absence of commuting to and from the workplace. Alcohol consumption increased in a quarter of respondents, possibly related to stress.

Research has shown adverse psychosocial changes during the COVID-19 pandemic, with higher levels of anxiety, depression, PTSD, and stress observed among the general population in multiple countries.^15^ In the U.S., a survey conducted from March through June 2020 found that distress increased as the pandemic first emerged in the U.S., including increases in anxiety and depression.^67^ Literature has suggested psychosocial support to be critical to mitigating these adverse changes, as individuals reporting having no psychosocial support were more vulnerable to mental health problems during the COVID-19 pandemic.^68^ These observations align with our findings from employees across the U.S. and suggest employers might need to make concerted efforts to assist employees in dealing with mental health issues as more individuals return to the office. Notably, our observations suggest that the focus of employers may need to be most concentrated on those who have worked the least amount of time at home during COVID-19.

Our behavioral outcome observations were also largely consistent with previous research on the COVID-19 pandemic. Increased non-work technology usage^14^ and screen time^69^ have been seen in previous studies. Interestingly, we saw a slight increase in hours of sleep per night among employees >75% WFH each week, but not among employees ≥ 25% WFH each week. Despite the increased sleep duration observed among some, other research during the COVID-19 pandemic has suggested individuals’ sleep quality has been poorer.^14^ Reductions in physical activity were also observed in existing research.^69^ More investigation of these phenomena, as well as changes in alcohol consumption, may be warranted. We believe our ongoing follow-up CAPTURE Surveys will provide more insight.

A secondary aim of CAPTURE was to assess personal SARS-CoV-2 transmission prevention behaviors taken at the workplace to mitigate the spread of COVID-19. While wearing a mask, washing hands regularly, physically distancing from coworkers, and monitoring symptoms were reported in high percentages, wearing gloves and disinfecting surfaces were not as common. This may be explained by the changing viewpoints of SARS-CoV-2 virus spread and associated mitigation strategy guidance set forth by the CDC^70^ and the World Health Organization’s (WHO)^71^ suggesting that surface-based transmission is less common. Importantly, employees reported their companies to be highly supportive of SARS-CoV-2 prevention behavior engagement. Indeed, many companies provided COVID-19 prevention training for their employees, which may have also aided in the high percentage of prevention behaviors reported.

Study limitations should be noted. First, participants were a convenience sample. Over 200 companies were contacted, but the companies that did partake in the survey had previously worked with, or were connected to, the research institutions. This convenience sample skewed the educational status of participants, with more than half of participants having a graduate degree. Second, while we asked about job status, we did not ask if employees identified as essential or non-essential workers due to the multidimensional nature of the companies. Future questionnaires should include perceptions of essential worker status or have a defined strategy to determine if employees are considered essential or non-essential workers. Finally, this was a cross-sectional analysis of a prospective survey. The questions asking about outcomes and perceptions “Before COVID-19” were therefore retrospective and subject to recall bias. However, we have future papers planned to describe prospective observations related to our ongoing follow-up CAPTURE Surveys. The survey was subjective and only asked about perceptions, feelings, and behaviors. Objective measures should be considered for future studies when feasible.

Nevertheless, the study had its strengths including: (1) a large sample size; (2) a wide range of WFH percentages; and (3) a sample of non-healthcare employees. These strengths provided unique insight into psychosocial and behavioral responses of employees working outside of the healthcare workforce during COVID-19. The increases in stress, anxiety, fatigue, and feeling unsafe due to work-related duties, as well as adverse changes in health behaviors (e.g., increased non-work screen time, decreased physical activity), suggest companies might consider how to support their employees as employees begin to return to the office in greater numbers.

## Supporting information

Appendix A: CAPTURE Baseline Survey

## Data Availability

Our data are available upon specific request to the corresponding author.

## Acknowledgements

The authors thank the staff of the Well Living Lab for their support. We acknowledge Qingyang Liu for her helpful guidance with analyzing the data, Laura Kjarland for assistance with survey deployment, and Austin Hoeg for his assistance with the project.

## References

1. University JH. Johns Hopkins Coronavirus Resource Center. https://coronavirus.jhu.edu/map.html Accessed May 11, 2020.

2. WHO. WHO director-general’s opening remarks at the media briefing on COVID-19 - 11 March 2020. https://www.who.int/dg/speeches/detail/who-director-general-s-opening-remarks-at-the-media-briefing-on-covid-1911-march-2020.

3. Richardson S, Hirsch J, Narasimhan M, et al. Presenting characteristics, comorbidities, and outcomes among 5700 patients hospitalized with COVID-19 in the New York City area. JAMA 2020;323(20):2052–2059.

4. He X, Lau E, Wu P, et al. Temporal dynamics in viral shedding and transmissibility of COVID-19. Nature Medicine 2020;26:672–675.

5. West R, Michie S, Rubin GJ, Amlot R. Applying principles of behaviour change to reduce SARS-CoV-2 transmission. Nature Human Behavior 2020;4:451–459.

6. Lake MA. What we know so far: COVID-19 current clinical knowledge and research. Clin Med (Lond) 2020;20(2):124–127.

7. Van Bavel JJ, Baicker K, Boggio PS, et al. Using social and behavioural science to support COVID-19 pandemic response. Nature Human Behavior 2020;4:460–471.

8. Zhang J, Lu H, Zeng H, et al. The differential psychological distress of populations affected by the COVID-19 pandemic. Brain, Behavior, and Immunity In Press: DOI: 10.1016/j.bbi.2020.03.007.

9. Holmes E, O’Connor R, Perry V, et al. Multidisciplinary research priorities for the COVID-19 pandemic: a call for action for mental health science. The Lancet In Press:DOI: 10.1016/S2215-0366(20)30168-1.

10. Dubey S, Biswas P, Ghosh R, et al. Psychosocial impact of COVID-19. Diabetes & Metabolic Syndrome: Clinical Research & Reviews 2020;14:779–788.

11. Wang C, Pan R, Wan X, et al. Immediate psychological responses and associated factors during the initial stage of the 2019 coronavirus disease (COVID-19) epidemic among the general population in China. International Journal of Environmental Research and Public Health 2020;17:doi: 10.3390/ijerph17051729.

12. Cao W, Fang Z, Hou G, et al. The psychological impact of the COVID-19 pandemic on college students in China. Psychiatry Research 2020;287:doi: 10.1016/j.psychres.2020.112934.

13. Latkin CA, Dayton L, Moran M, Strickland JC, Collins K. Behavioral and psychosocial factors associated with COVID-19 skepticism in the United States. Curr Psychol 2021:1–9.

14. Ammar A, Trabelsi K, Brach M, et al. Effects of home confinement on mental health and lifestyle behaviours during the COVID-19 outbreak: insights from the ECLB-COVID19 multicentre study. Biol Sport 2021;38(1):9–21.

15. Hossain MM, Tasnim S, Sultana A, et al. Epidemiology of mental health problems in COVID-19: a review. F1000Res 2020;9:636.

16. Hossain MM, Sultana A, Purohit N. Mental health outcomes of quarantine and isolation for infection prevention: a systematic umbrella review of the global evidence. Epidemiol Health 2020;42:e2020038.

17. Liu J, Liao X, Qian S, et al. Community transmission of severe acute respiratory syndrome coronavirus 2, Shenzhen, China, 2020. Emerging Infectious Diseases 2020: DOI: 10.3201/eid2606.200239.

18. Chan J-W, Yuan S, Kok K-H, et al. A familial cluster of pneumonia associated with the 2019 novel coronavirus indicating person-to-person transmission: a study of a family cluster. The Lancet 2020;395(10223):P514–523.

19. Li Q, Guan X, Wu P, et al. Early transmission dynamics in Wuhan, China, of novel coronavirus-infected pneumonia. New England Journal of Medicine 2020;382.

20. Huang C, Wang Y, Li X, et al. Clinical features of patients infected with 2019 novel coronavirus in Wuhan, China. The Lancet 2020;395(10223):P497–506.

21. Burke R, Midgley C, Dratch A, et al. Active monitoring of persons exposed to patients with confirmed COVID-19--United States, January-February 2020. Morbidity and Mortality Weekly Report 2020;69(9).

22. WHO. Report of the WHO-China joint mission on coronavirus disease 2019 (COVID-19). Geneva, Switzerland: World Health Organization, 2020.

23. Ong S, Tan Y, Chia P, et al. Air, surface environmental, and personal protective equipment contamination by severe acute respiratory syndrome coronavirus 2 (SARS-CoV-2) from a symptomatic patient. JAMA 2020;323(16):1610–1612.

24. Ai Z, Melikov A. Airborne spread of expiratory droplet nuclei between the occupants of indoor environments: a review. Indoor Air 2018;28(4):doi: 10.1111/ina.12465.

25. Dietz L, Horve P, Coil D, et al. 2019 novel coronavirus (COVID-19) pandemic: built environment considerations to reduce transmission. mSystems 2020;5:doi: 10.1128/mSystems.00245-20.

26. van Doremalen N, Bushmaker T, Morris D, et al. Aerosol and surface stability of SARS-CoV-2 as compared with SARS-CoV-1. New England Journal of Medicine 2020;382(16):1564–1567.

27. Liu Y, Ning Z, Chen Y, et al. Aerodynamic Analysis of SARS-CoV-2 in two Wuhan hospitals. Nature 2020: DOI: 10.1038/s41586-020-2271-3(2020).

28. Schoen L. Guidance for building operations during the COVID-19 pandemic. ASHRAE Journal. Atlanta, GA: American Society of Heating Refrigerating and Air-Conditioning Engineers, 2020.

29. Otter J, Donskey C, Yezli S, et al. Transmission of SARS and MERS coronaviruses and influenza virus in healthcare settings: the possible role of dry surface contamination. Journal of Hospital Infection 2016;92:235–250.

30. Bahl P, Doolan C, de Silva C, et al. Airborne or droplet precautions for health workers treating coronavirus disease 2019. Journal of Infectious Diseases 2020:DOI: 10.1093/infdis/jiaa189.

31. Vejerano E, Marr L. Physio-chemical characteristics of evaporating respiratory fluid droplets. Journal of the Royal Society Interface 2018:DOI: 10.1098/rsif.2017.0939.

32. Yang S, Lee G, Chen C-M, Wu C-C, Yu K-P. The size and concentration of droplets generated by coughing in human subjects. J Aerosol Med 2007;20(4):484–494.

33. Fennelly K, Martyny J, Fulton K, et al. Cough-generated aerosols of mycobacterium tuberculosis: A new method to study infectiousness. Am J Respir Crit Care Med 2004;169(5):604–609.

34. Gupta J, Lin C-H, Chen Q. Flow dynamics and characterization of a cough. Indoor Air 2009;19(6):517–525.

35. Xie X, Li Y, Sun H, Liu L. Exhaled droplets due to talking and coughing. J R Soc Interface 2009;6:doi: 10.1098/rsif.2009.0388.focus.

36. Zhu S, Kato S, Yang J-H. Study on transport characteristics of saliva droplets produced by couching in a calm indoor environment. Building and Environment 2006;41(12):1691–1702.

37. Louden R, Roberts R. Droplet expulsion from the respiratory tract. American Review of Respiratory Disease 1967;95(3):435–442.

38. Duguid J. The size and duration of air-carriage of respiratory droplets and droplet-nuclei. Epidemiol Infect 1946;44(6):471–479.

39. Lindsley W, Pearce T, Hudnall J, et al. Quantity and size distribution of cough-generated aerosol particles produced by influenza patients during and after illness. J Occup Environ Hyg 2012;9(7):443–449.

40. Lindsley W, Reynolds J, Szalajda J, Noti J, Beezhold D. A cough aerosol simulator for the study of disease transmission by human cough-generated aerosols. Aerosol Science and Technology 2013;47(8):937–944.

41. Lindsley W, Blachere F, Beezhold D, et al. Viable influenza A virus in airborne particles expelled during coughs versus exhalations. Influenza Other Respir Viruses 2016;10(5):404–413.

42. Bourouiba L, Dehandschoewercker E, Bush J. Violent expiratory events: on coughing and sneezing. Journal of Fluid Dynamics 2014;745:537–563.

43. Zayas G, Chiang M, Wong E, et al. Cough aerosol in healthy participants: fundamental knowledge to optimize droplet-spread infectious respiratory disease management. BMC Pulmonary Medicine 2012;11:doi: 10.1186/1471-2466-12-11.

44. Scharfman B, Techet A, Bush J, Bourouiba L. Visualization of sneeze ejecta: steps of fluid fragmentation leading to respiratory droplets. Experiments in Fluids 2016;57(2):24.

45. Han Z, Weng W, Huang Q. Characterizations of particle size distribution of the droplets exhaled by sneeze. J R Soc Interface 2013;10(88):doi: 10.1098/rsif.2013.0560.

46. Morawska L, Johnson G, Ristovski Z, et al. Size distribution and sites of origin of droplets expelled from the human respiratory tract during expiratory activities. Journal of Aerosol Science 2009;40(3):256–269.

47. Edwards D, Man J, Brand P, et al. Inhaling to mitigate exhaled bioaerosols. PNAS 2004;101(50):17383–17388.

48. Papineni R, Rosenthal F. The size distribution of droplets in the exhaled breath of healthy human subjects. Journal of Aerosol Medicine 1997;10(2):105–116.

49. Fabian P, McDevitt J, DeHaan W, et al. Influenza virus in human exhaled breath: an observational study. PLoS One 2008;3(7):doi: 10.1371/journal.pone.0002691.

50. Asadi S, Wexler A, Cappa C, et al. Aerosol emission and superemission during human speech increase with voice loudness. Scientific Reports 2019;9(1):1–10.

51. Gupta J, Lin C-H, Chen Q. Characterizing exhaled airflow from breathing and talking. Indoor Air 2010;20(1):31–39.

52. Stadnytskyi V, Bax C, Bax A, Anfinrud P. The airborne lifetime of small speech droplets and their potential importance in SARS-CoV-2 transmission. PNAS 2020;117(22):11875–11877.

53. Chu D, Akl E, Duda S, et al. Physical distancing, face masks, and eye protection to prevention person-to-person transmission of SARS-CoV-2 and COVID-19: A systematic review and meta-analysis. Lancet 2020;395:1973–1987.

54. Rabenau H, Kampf G, Cinatl J, Doerr H. Efficacy of various disinfectants against SARS coronavirus. Journal of Hospital Infection 2005;61:107–111.

55. Sattar S, Springthorpe V, Karim Y, Loro P. Chemical disinfection of non-porous inanimate surfaces experimentally contaminated with four human pathogenic viruses. Epidem Inf 1989;102:493–505.

56. Wood A, Payne D. The action of three antiseptics/disinfectants against enveloped and non-enveloped viruses. Journal of Hospital Infection 1998;38:283–295.

57. Kariwa H, Fujii N, Takashima I. Inactivation of SARS coronavirus by means of povidone-iodine, physical conditions and chemical reagents. Dermatology 2006;212:119–123.

58. Omidbakhsh N, Sattar S. Broad-spectrum microbicidal activity, toxicologic assessment, and materials compatibility of a new generation of accelerated hydrogen peroxide-based environmental surface disinfectant. Am J Infect Control 2006;34:251–257.

59. Eggers M, Eickmann M, Zorn J. Rapid and effective virucidal activity of povidone-iodine products against Middle East respiratory syndrome coronavirus (MERS-CoV) and modified vaccinia virus ankara (MVA). Infect Dis Ther 2015;4:491–501.

60. CDC. Coronavirus disease 2019: businesses & workplaces-plan, prepare, and respond. Atlanta, GA: Centers for Disease Control and Prevention, 2020.

61. Friedman GD, Cutter GR, Donahue RP, et al. CARDIA: Study Design, Recruitment, and Some Characteristics of the Examined Subjects. J Clin Epidemiol 1988;41(11):1105–1116.

62. Pereira M, Mullane S, Toledo MJ, et al. Efficacy of the ‘Stand and Move at Work’ multicomponent workplace intervention to reduce sedentary time and improve cardiometabolic risk: a group randomized clinical trial. International Journal of Behavioral Nutrition and Physical Activity 2020;17(133).

63. (NIH) NIoH. COVID-19 Questionnaire - Adult Primary Version. In: (ECHO) EioCHO, ed, 2020.

64. Manning KJ, Steffens, D.C., Grasso, D.J., Briggs-Gowan, M.J., Ford, J.D., & Carter, A.S. The Epidemic - Pandemic Impacts Inventory Geriatric Adaptation (EPII-G). University of Connecticut School of Medicine, 2020.

65. Hughes ME, Waite LJ, Hawkley LC, Cacioppo JT. A Short Scale for Measuring Loneliness in Large Surveys: Results from Two Population-Based Studies. Res Aging 2004;26:655–672.

66. Conway LG III, Woodard, S. R., & Zubrod, A. Social Psychological Measurements of COVID-19: Coronavirus Perceived Threat, Government Response, Impacts, and Experiences Questionnaires.. https://psyarxiv.com/z2x9a/.

67. Daly M, Robinson E. Psychological distress and adaptation to the COVID-19 crisis in the United States. J Psychiatr Res 2021;136:603–609.

68. Lei L, Huang X, Zhang S, et al. Comparison of Prevalence and Associated Factors of Anxiety and Depression Among People Affected by versus People Unaffected by Quarantine During the COVID-19 Epidemic in Southwestern China. Med Sci Monit 2020;26:e924609.

69. Meyer J, McDowell C, Lansing J, et al. Changes in Physical Activity and Sedentary Behavior in Response to COVID-19 and Their Associations with Mental Health in 3052 US Adults. Int J Environ Res Public Health 2020;17(18).

70. Prevention CfDCa. COVID-19. https://www.cdc.gov/coronavirus/2019-ncov/index.html Accessed May 7, 2021.

71. (WHO) WHO. Coronavirus disease (COVID-19): How is it transmitted? World Health Organization (WHO), 13 December 2020.

