## Appendix A: CAPTURE Baseline Survey for "Psychosocial and Behavioral Responses and SARS-CoV-2 Transmission Prevention Behaviors while Working during the COVID-19 Pandemic"

**Research Study:** Working During the COVID-19 Pandemic: Characterizing Awareness of SARSCoV-2 PrevenTion and Understanding Responses and Experiences (CAPTURE) Survey

Please click the link to read the Consent Form and sign below.

I have read the Consent Form and agree to participate in this research study.

Are you aged 18 years or older?

- ◯ Yes
- ◯ No

*Skip To: End of Survey If Are you aged 18 years or older? = No*

---

Please enter the email address* at which you received this survey:

**Please note that we will only ask for your email so that we may contact you with an invitation to complete the survey at 3-, 6-, and 12-month timepoints. All emails are maintained in a secure, password-protected database that will only be used to contact you at the future timepoints. Emails will be deidentified (stripped) from all responses prior to data analysis. Please see consent form for more information.*

________________________________________________________________

Please state the name of the company you work for.

________________________________________________________________

In which state do you currently work in?

▼ Alabama (1) ... I do not reside in the United States (53)

*Skip To: End of Survey If 50 States, D.C. and Puerto Rico = I do not reside in the United States*

**Working During the COVID-19 Pandemic: The CAPTURE Survey**

*Characterizing Awareness of SARS-CoV-2 PrevenTion and Understanding Responses and Experiences*

Thank you for your willingness to complete this survey! It should take no longer than 20 minutes to complete. As you will see, some questions are personal in nature, and others assess work-related behaviors as they relate to the current coronavirus disease-19 (**COVID-19**) pandemic. Severe acute respiratory syndrome coronavirus 2 (**SARS-CoV-2**), the virus causing COVID-19, refers to the illness caused by the novel coronavirus that was first identified in 2019.

We realize there are many different opinions on both the severity of, and responses to, the pandemic. Please know that **your answers to these questions are strictly confidential** and study results will only be presented in summary form. With that, we ask you to be honest in your responses.

When answering these questions, please only think about your work at [Company].

We very much appreciate your time and honesty.

If you have any questions while taking this study, please email the study coordinator.

Study contacts:
**Well Living Lab:** Araliya Senerat at.
**University of Minnesota:** Sarah Rydell at.

---

Are you an employee of [Company] and currently employed?

○ Yes

○ No

*Skip To: End of Survey If Are you an employee of ${q://QID393/ChoiceTextEntryValue} and currently employed? = No*

On average, what percentage (%) of time do you spend indoors working in your current role at [Company]?

| | 0 10 20 30 40 50 60 70 80 90 100 |
|---|---|
| % of time indoors () | |

*Skip To: End of Survey If On average, what percentage (%) of time do you spend indoors working in your current role at ... [ % of time indoors ] < 50*

How many hours per week, on average, do you work at this job?

- ◯ 0 hours/wk
- ◯ 1-8 hours
- ◯ 9-16
- ◯ 17-24
- ◯ 25-32
- ◯ 33-40
- ◯ 41-60
- ◯ 61 or more
- ◯ Prefer not to answer

On average, how many hours per week do you <u>currently</u> have face-to-face, in-person interactions with either your coworkers or the public <u>while completing your job-related duties</u>?

- ◯ 0 hours/wk
- ◯ 1-8 hours
- ◯ 9-16
- ◯ 17-24
- ◯ 25-32
- ◯ 33-40
- ◯ 41-60
- ◯ 61 or more
- ◯ Prefer not to say

Please choose the category that best describes your main job. If none of the categories fits you exactly, please respond with the closest category to your experience. (Select only one.)

- ◯ Executive, administrator, or senior manager (e.g., CEO, sales, VP, plant manager)
- ◯ Professional (e.g., engineer, accountant, systems analyst)
- ◯ Technical support (e.g., lab technician, legal assistant, computer programmer)
- ◯ Sales (e.g., sales representative, stockbroker, retail sales)
- ◯ Clerical and administrative support (e.g., secretary, billing clerk, office supervisor)
- ◯ Service occupation (e.g., security officer, carpenter, machinist)
- ◯ Chemical/Production Operator (e.g., shift supervisors and hourly employees)
- ◯ Laborer (e.g., truck driver, construction worker)
- ◯ Food industry service occupation (e.g. server, chef, cook)
- ◯ Other ________________________________________________
- ◯ Prefer not to answer

**Before the COVID-19 pandemic**, what percentage of your work hours (weekly average) were you working at home?

| | Prefer not to answer<br>0 10 20 30 40 50 60 70 80 90 100 |
|---|---|
| Percentage (%) at home () | |

Now, **during the current COVID-19 pandemic**, what percentage of your work hours (weekly average) are you working at home?

| | Prefer not to answer<br>0 10 20 30 40 50 60 70 80 90 100 |
|---|---|
| Percentage (%) at home () | |

At [Company], are you now:

- ○ working full time, same hours as before pandemic
- ○ working full time, different hours than before pandemic
- ○ working part-time, same hours as before pandemic
- ○ working part-time,  different hours than before pandemic
- ○ retired
- ○ unemployed
- ○ permanently laid off
- ○ furloughed/furloughed previously (or temporarily laid off)

Please indicate whether any of these situations apply to you (check all that apply):

- ☐ Working remotely or from home
- ☐ Had to get a second job
- ☐ Working a second job, same hours as before pandemic
- ☐ Working a second job, different hours than before pandemic
- ☐ At increased risk of getting COVID-19 in your job

Please indicate the extent to which you view the COVID-19 outbreak as having either a positive or negative impact on your work.

- ◯ Extremely negative
- ◯ Moderately negative
- ◯ Somewhat negative
- ◯ No impact
- ◯ Slightly positive
- ◯ Moderately positive
- ◯ Extremely positive
- ◯ Prefer not to answer

**As a direct result of the COVID pandemic:**

*Display This Question:*

*If At [Company], are you now: = working part-time,  different hours than before pandemic*

You said you were working part time at [Company], but different hours before the pandemic. Are you working more hours or fewer hours?

- ◯ More hours
- ◯ Fewer hours
- ◯ Prefer not to answer

*Display This Question:*

*If Please indicate whether any of these situations apply to you (check all that apply): = Working a second job, different hours than before pandemic*

You said you were working part time, but different hours at your second job before the pandemic. Are you working more hours or fewer hours?

- ◯ More hours

○ Fewer hours

○ Prefer not to answer

Have you lost health insurance or other coverage for medical care?

○ Yes

○ No

○ Don't Know

○ Prefer not to answer

**In the past month**, I have taken the following precautions **at my company's workplace**:

| | Never | Rarely | Sometimes | Often | Always | Not Applicable |
|---|---|---|---|---|---|---|
| Worn a mask of any type | ○ | ○ | ○ | ○ | ○ | ○ |
| Worn gloves | ○ | ○ | ○ | ○ | ○ | ○ |
| Washed my hands regularly | ○ | ○ | ○ | ○ | ○ | ○ |
| Physically distanced from coworkers or public | ○ | ○ | ○ | ○ | ○ | ○ |
| Disinfected surfaces at which I primarily work | ○ | ○ | ○ | ○ | ○ | ○ |
| Monitored symptoms prior to work (e.g., measuring temperature) | ○ | ○ | ○ | ○ | ○ | ○ |
| Other, please describe: | ○ | ○ | ○ | ○ | ○ | ○ |

**In the past month,** how often have you observed your co-workers taking the following precautions **at your company's workplace**?

| | Never | Rarely | Sometimes | Often | Always | Not Applicable |
|---|---|---|---|---|---|---|
| Worn a mask of any type | | | | | | |
| Worn gloves | | | | | | |
| Washed their hands regularly | | | | | | |
| Physically distanced from coworkers or public | | | | | | |
| Disinfected surfaces at which they primarily work | | | | | | |
| Other, please describe: | | | | | | |

**During the COVID-19 pandemic**, [Company] has **provided**:

| | Never | Rarely | Sometimes | Often | Always | Not Applicable |
|---|---|---|---|---|---|---|
| N95 masks | ○ | ○ | ○ | ○ | ○ | ○ |
| Surgical masks | ○ | ○ | ○ | ○ | ○ | ○ |
| Cloth masks | ○ | ○ | ○ | ○ | ○ | ○ |
| Gloves | ○ | ○ | ○ | ○ | ○ | ○ |
| Hand sanitizer | ○ | ○ | ○ | ○ | ○ | ○ |
| Hand washing instructions | ○ | ○ | ○ | ○ | ○ | ○ |
| Physical distancing instructions | ○ | ○ | ○ | ○ | ○ | ○ |
| Cleaning/disinfecting products for surfaces | ○ | ○ | ○ | ○ | ○ | ○ |
| Tools to monitor symptoms prior to work (e.g., thermometer to measure temperature) | ○ | ○ | ○ | ○ | ○ | ○ |
| Other, please describe: | ○ | ○ | ○ | ○ | ○ | ○ |

**During the COVID-19 pandemic**, [Company] has **promoted**:

| | Never | Rarely | Sometimes | Often | Always | Not Applicable |
|---|---|---|---|---|---|---|
| N95 masks | ○ | ○ | ○ | ○ | ○ | ○ |
| Surgical masks | ○ | ○ | ○ | ○ | ○ | ○ |
| Cloth masks | ○ | ○ | ○ | ○ | ○ | ○ |
| Gloves | ○ | ○ | ○ | ○ | ○ | ○ |
| Hand sanitizer | ○ | ○ | ○ | ○ | ○ | ○ |
| Hand washing | ○ | ○ | ○ | ○ | ○ | ○ |
| Physical distancing | ○ | ○ | ○ | ○ | ○ | ○ |
| Surface cleaning/disinfecting | ○ | ○ | ○ | ○ | ○ | ○ |
| Monitoring of symptoms prior to work (e.g., thermometer to measure temperature) | ○ | ○ | ○ | ○ | ○ | ○ |
| Other, please describe: | ○ | ○ | ○ | ○ | ○ | ○ |

What best describes the COVID-19 prevention training that your employer has provided?

- ☐ ⊗ None
- ☐ Web training
- ☐ In-person training
- ☐ Reading materials
- ☐ Other, please describe_______________________________________________
- ☐ Prefer not to answer

How important do you think the following practices are in the prevention of the spread of COVID-19?:

| | Not important | Somewhat important | Moderately important | Very important | Prefer not to answer |
|---|---|---|---|---|---|
| Wearing a mask | ○ | ○ | ○ | ○ | ○ |
| Wearing gloves | ○ | ○ | ○ | ○ | ○ |
| Handwashing | ○ | ○ | ○ | ○ | ○ |
| Physical distancing | ○ | ○ | ○ | ○ | ○ |
| Disinfecting surface | ○ | ○ | ○ | ○ | ○ |
| Other, please describe: | ○ | ○ | ○ | ○ | ○ |

How much did you experience the following feelings due to your work-related duties **before COVID-19** and **now, during COVID-19**?

| | Before COVID-19 | | | | | | During COVID-19 | | | | | |
|---|---|---|---|---|---|---|---|---|---|---|---|---|
| | Never | Rarely | Moderately | Quite a Bit | All the time | Prefer not to answer | Never | Rarely | Moderately | Quite a Bit | All the time | Prefer not to answer |
| Stress | | | | | | | | | | | | |
| Anxiety | | | | | | | | | | | | |
| Fatigue | | | | | | | | | | | | |
| Feeling unsafe | | | | | | | | | | | | |

How much do you agree or disagree with the following statements?

| | Strongly Disagree | Disagree | Neutral | Agree | Strongly Agree |
|---|---|---|---|---|---|
| Thinking about the coronavirus (COVID-19) makes me feel threatened | ○ | ○ | ○ | ○ | ○ |
| I am afraid of the coronavirus (COVID-19) | ○ | ○ | ○ | ○ | ○ |
| I am stressed around other people because I worry I'll catch the coronavirus (COVID-19) | ○ | ○ | ○ | ○ | ○ |

How would you describe your level of **productivity** in your job **before COVID-19** and **now, during COVID-19**?

| | Low | Slightly below average | Average | Slightly above average | High | Prefer not to answer |
|---|---|---|---|---|---|---|
| **Before COVID-19** | ○ | ○ | ○ | ○ | ○ | ○ |
| **During COVID-19** | ○ | ○ | ○ | ○ | ○ | ○ |

How often have you felt the following **at work before COVID-19** and **now, during COVID-19?**

| | **Before COVID-19** | | | | **During COVID-19** | | | |
|---|---|---|---|---|---|---|---|---|
| | Hardly ever | Some of the time | Often | Prefer not to answer | Hardly ever | Some of the time | Often | Prefer not to answer |
| A lack of companionship | ○ | ○ | ○ | ○ | ○ | ○ | ○ | ○ |
| A feeling of being left out | ○ | ○ | ○ | ○ | ○ | ○ | ○ | ○ |
| A feeling of being isolated from others | ○ | ○ | ○ | ○ | ○ | ○ | ○ | ○ |

How many **minutes** per day did you spend physically active (i.e., walking, jogging, swimming, gardening, house-chores) **before COVID-19** and, now, **during COVID-19**?

| | 0-30 minutes per day | 30-60 minutes per day | 60-90 minutes per day | 90-120 minutes per day | >120 minutes per day | Prefer not to answer |
|---|---|---|---|---|---|---|
| **Before COVID-19** | ○ | ○ | ○ | ○ | ○ | ○ |
| **During COVID-19** | ○ | ○ | ○ | ○ | ○ | ○ |

How many **hours** per day did you watch television, use the computer for non-work, utilize your phone for entertainment, play video games **before COVID-19** and, now, **during COVID-19**?

| | 0-1 hour per day | 1-2 hours per day | 2-4 hours per day | >5 hours per day | Prefer not to answer |
|---|---|---|---|---|---|
| **Before COVID-19** | ○ | ○ | ○ | ○ | ○ |
| **During COVID-19** | ○ | ○ | ○ | ○ | ○ |

How many **hours** per night of sleep did you get **before COVID-19** and, now, **during COVID-19**?

| | <6 hours per night | 7 hours per night | 8 hours per night | 9 hours per night | >10 hours per night | Prefer not to answer |
|---|---|---|---|---|---|---|
| **Before COVID-19** | ○ | ○ | ○ | ○ | ○ | ○ |
| **During COVID-19** | ○ | ○ | ○ | ○ | ○ | ○ |

How often did you have any kind of drink containing alcohol **on average each week before COVID-19** and, now, **during COVID-19**?
By a drink we mean half an ounce of absolute alcohol (e.g. a 12 ounce can or glass of beer or cooler, a 5 ounce glass of wine, or a drink containing 1 shot of liquor).
Choose only one.

| | 0 drinks/ week | 1-3 drinks/ week | 4-6 drinks/ week | 7-9 drinks/ week | 10-12 drinks/ week | 13-15 drinks/ week | 16-18 drinks/ week | 19-21 drinks/ week | >21 drinks/ week | Prefer not to answer |
|---|---|---|---|---|---|---|---|---|---|---|
| **Before COVID-19** | | | | | | | | | | |
| **During COVID-19** | | | | | | | | | | |

How would you categorize your use of the following tobacco/nicotine products **before COVID-19** and **now, during COVID-19**?

| | **Before COVID-19** | | | | | **During COVID-19** | | | | |
|---|---|---|---|---|---|---|---|---|---|---|
| | Increased | Decreased | Stayed the same | Never used | Prefer not to answer | Increased | Decreased | Stayed the same | Never used | Prefer not to answer |

| | | | | | | | | | | |
|---|---|---|---|---|---|---|---|---|---|---|
| Cigarettes | ○ | ○ | ○ | ○ | ○ | ○ | ○ | ○ | ○ | ○ |
| E-cigarettes | ○ | ○ | ○ | ○ | ○ | ○ | ○ | ○ | ○ | ○ |
| Pipe, cigars, or cigarillos | ○ | ○ | ○ | ○ | ○ | ○ | ○ | ○ | ○ | ○ |
| Smokeless tobacco (e.g. snuff, chewing tobacco, dip) | ○ | ○ | ○ | ○ | ○ | ○ | ○ | ○ | ○ | ○ |

How worried are you that you'll contract COVID-19 while at work?

- ○ Not at all worried
- ○ Somewhat worried
- ○ Moderately worried
- ○ Very worried
- ○ Prefer not to answer

How worried are you that you could be an asymptomatic carrier of COVID-19 and may be spreading it to other people **at your company's workplace**?

- ◯ Not at all worried
- ◯ Somewhat worried
- ◯ Moderately worried
- ◯ Very worried
- ◯ Prefer not to answer

To help us better understand the health of all study participants during the COVID-19 pandemic, we would like to ask you questions about your possible exposure to this new virus. We use the term COVID-19 to refer to the illness caused by the novel coronavirus that was first identified in 2019. This virus is also called SARS-CoV-2.

Below is a list of symptoms that may be related to COVID-19. Some of these may also occur with other conditions such as allergies, colds and flu or when taking certain medications.

Have you had any of these symptoms for longer than several hours or more than is usual for you, **since March 2020**?

| | No | Yes | Prefer not to answer |
|---|---|---|---|
| Fever (≥100.4°F) | ○ | ○ | ○ |
| Persistent cough | ○ | ○ | ○ |
| Unusual shortness of breath or difficulty breathing | ○ | ○ | ○ |
| Chills or sweats | ○ | ○ | ○ |
| Headache | ○ | ○ | ○ |
| Sore throat | ○ | ○ | ○ |
| Unusually hoarse | ○ | ○ | ○ |
| Loss of smell | ○ | ○ | ○ |
| Loss of taste | ○ | ○ | ○ |
| Chest pain/tightness | ○ | ○ | ○ |
| Muscle aches | ○ | ○ | ○ |
| Abdominal pain | ○ | ○ | ○ |
| Diarrhea | ○ | ○ | ○ |
| Confusion | ○ | ○ | ○ |
| Malaise or general feeling of illness, discomfort or uneasiness | ○ | ○ | ○ |
| Unusual fatigue | ○ | ○ | ○ |

*Skip To: Q35 If Have you had any of these symptoms for longer than several hours or more than is usual for you, s... = No*

*Carry Forward Selected Choices from "Have you had any of these symptoms for longer than several hours or more than is usual for you, since March 2020?"*

If **YES**, how severe was this symptom?

| | Mild | Moderate | Severe | Prefer not to answer |
|---|---|---|---|---|
| Fever (≥100.4°F) | ◯ | ◯ | ◯ | ◯ |
| Persistent cough | ◯ | ◯ | ◯ | ◯ |
| Unusual shortness of breath or difficulty breathing | ◯ | ◯ | ◯ | ◯ |
| Chills or sweats | ◯ | ◯ | ◯ | ◯ |
| Headache | ◯ | ◯ | ◯ | ◯ |
| Sore throat | ◯ | ◯ | ◯ | ◯ |
| Unusually hoarse | ◯ | ◯ | ◯ | ◯ |
| Loss of smell | ◯ | ◯ | ◯ | ◯ |
| Loss of taste | ◯ | ◯ | ◯ | ◯ |
| Chest pain/tightness | ◯ | ◯ | ◯ | ◯ |
| Muscle aches | ◯ | ◯ | ◯ | ◯ |
| Abdominal pain | ◯ | ◯ | ◯ | ◯ |
| Diarrhea | ◯ | ◯ | ◯ | ◯ |
| Confusion | ◯ | ◯ | ◯ | ◯ |
| Malaise or general feeling of illness, discomfort or uneasiness | ◯ | ◯ | ◯ | ◯ |
| Unusual fatigue | ◯ | ◯ | ◯ | ◯ |

Which of the following statements apply to you?

- ○ I know I don't have or have not had COVID-19 due to having undergone testing.
- ○ I do not think I had a COVID-19 infection and/or have had no symptoms.
- ○ I suspected that I had a COVID-19 infection but I never sought medical care.
- ○ I called my health care provider because I thought I might have a COVID-19 infection and I was told to stay home (quarantine).
- ○ I went to a clinic, emergency room or hospital because I had symptoms that might be from COVID-19.
- ○ Prefer not to answer

Have you had the nasal swab or spit test (also known as PCR test) for the virus that causes COVID-19? (***Mark all that apply***)

- ☐ No, I never tried to get tested
- ☐ No, I tried to get tested but was not able to
- ☐ Yes, and I am waiting for the results
- ☐ Yes, and the test showed that I did not have it ("negative" test)
- ☐ Yes, and the test showed that I did have it ("positive" test)
- ☐ ⊗ Prefer not to answer

Have you had a blood test (also known as an antibody test) to see whether you already had the COVID-19 virus ("serology")? ***(Mark all that apply)***

- ☐ No, I never tried to get tested
- ☐ No, I tried to get tested but was not able to
- ☐ Yes, and I am waiting for the results
- ☐ Yes, and the test showed that I did not have it ("negative" test)
- ☐ Yes, and the test showed that I did have it ("positive" test)
- ☐ ⊗ Prefer not to answer

In what ways has the COVID-19 outbreak affected your overall healthcare (e.g., primary care provider, dentist, eye doctor, therapist, etc.)? ***(Mark all that apply)***

- ☐ I did not go to **some** healthcare appointments because I was concerned about entering my healthcare providers' office
- ☐ I did not go to **any** healthcare appointments because I was concerned about entering my healthcare providers' office
- ☐ My healthcare provider(s) canceled appointments
- ☐ My healthcare provider(s) changed to phone or online visits
- ☐ My healthcare provider(s) told me to self-isolate or quarantine
- ☐ ⊗ None of these apply
- ☐ ⊗ Prefer not to answer

Which of the following behaviors have you **done less of** because of the COVID-19 outbreak? *(**Mark all that apply**)*

- ☐ In-person contact with people inside my home (that is, you are quarantined separately from one or more family or household members)
- ☐ In-person contact with family who live outside the home
- ☐ In-person contact with friends
- ☐ In-person contact with colleagues at work
- ☐ In-person events in the community, including religious events
- ☐ ⊗ None of these apply
- ☐ ⊗ Prefer not to answer

In general, would you say your health is:

- ○ Poor
- ○ Fair
- ○ Good
- ○ Very Good
- ○ Excellent
- ○ Prefer not to answer

Have you ever been told by a doctor that you have...

| | Yes | No | Not Sure | Prefer not to answer |
|---|---|---|---|---|
| Heart disease or angina | ○ | ○ | ○ | ○ |
| High blood pressure (hypertension) | ○ | ○ | ○ | ○ |
| Stroke | ○ | ○ | ○ | ○ |
| Diabetes (high blood sugar) | ○ | ○ | ○ | ○ |
| Cancer | ○ | ○ | ○ | ○ |
| High cholesterol | ○ | ○ | ○ | ○ |
| An anxiety disorder (e.g. GAD) | ○ | ○ | ○ | ○ |
| A mood disorder (e.g. depression) | ○ | ○ | ○ | ○ |

What is your gender?

○ Female

○ Male

○ Other/Non-binary

○ Prefer not to answer

**Demographic Information**

What is your current age?

________________________________________________________________

Are you Hispanic or Latino/Latina/Latinx?

- ◯ Yes
- ◯ No
- ◯ I prefer not to answer

Which of the following best describes you? (Check all that apply)

- ☐ Asian
- ☐ Black or African-American
- ☐ Hawaiian or Pacific Islander
- ☐ Native American or Alaskan Native
- ☐ Hispanic or Latino/Latina/Latinx
- ☐ White
- ☐ Other (please specify)
- ☐ I prefer not to answer

What is your current marital status?

- ◯ Single
- ◯ Married or partnered
- ◯ I prefer not to answer

What is the highest level of schooling you completed?

- ◯ Less than High School
- ◯ Obtained GED
- ◯ High School Graduate (diploma)
- ◯ Completed some college credit, but no degree
- ◯ Associate degree
- ◯ Bachelor's degree
- ◯ Master's, Professional, or Doctoral degree
- ◯ I prefer not to answer

Do you have any other thoughts or feelings regarding working during the COVID-19 pandemic that you do not feel might have been captured within this survey? If so, please describe.

________________________________________________________________
